# Detecting orientation of Brain MR scans using deep learning

**DOI:** 10.1101/2021.08.17.21262189

**Authors:** Chinmay Singhal, Nihit Gupta, Anouk Stein, Quan Zhou, Leon Chen, George Shih

## Abstract

There has been a steady escalation in the impact of Artificial Intelligence (AI) on Healthcare along with an increasing amount of progress being made in this field. While many entities are working on the development of significant deep learning models for the diagnosis of brain-related diseases, identifying precise images needed for model training and inference tasks is limited due to variation in DICOM fields which use free text to define things like series description, sequence and orientation [1]. Detecting the orientation of brain MR scans (Axial/Sagittal/Coronal) remains a challenge due to these variations caused by linguistic barriers, human errors and de-identification - essentially rendering the tags unreliable [2, 3, 4]. In this work, we propose a deep learning model that identifies the orientation of brain MR scans with near perfect accuracy.

## 1 Introduction

The utility of AI in the healthcare sector has been globally increasing. It’s current market size value in 2021 is USD 10.4 billion and is expected to expand to USD 120.2 billion by the end of 2028 [5]. Further the growth in the field of medical imaging and diagnostics can be attributed to the existence of a large amount of imaging data, availability of many AI systems to radiologists for managing diagnosis and treatment and the influx of a large number of startups in this segment [6]. In our work, we develop a solution for detecting the orientation of brain MR scans. Our solution aims to find its use in the management of large datasets, data annotations, and model testing to be used by various entities working in the area of developing healthcare AI.

The current works in this area suggest methods relying on the metadata of the DICOM files but the real question is how reliable is the metadata? The values of various attributes in the DICOM metadata are unreliable and inconsistent [2, 3, 4]. Automating the classification of the orientation plane of the brain MR scans thus becomes challenging. Given this context, we aim to develop a reliable solution that utilizes the image pixels for determining the orientation plane. In this article, we describe the method of using a deep learning model and includes the following parts:

- Annotating the series containing the DICOM files using the computation of Image Orientation Plane attribute’s value method and verifying the annotations.
- Pre-processing the pixel data of DICOM files for feature extraction.
- Using a pre-trained neural network to train and fine-tune the classifier.

Foremost, we present the usual and conventional methods of detecting the orientation of the Brain MR scans. After that, we describe the various aspects of the proposed solution, and then we finally present our results.

## 2 Background Information

Various works suggests retrieving the orientation plane from the “SeriesDescription” DICOM attribute as it is often specified in a series description. For example, “AX” in “AX T1 pre gd” means axial. However, orientation is not always included in the description. For example, retrieving the orientation plane from the series description,“T2-weighted trace” is not possible using this method. Current practice also suggests using the DICOM attribute Image Orientation Patient (0020,0037). The attribute values give the direction cosines of the first row and first column of an image with respect to the patient which could be further used to compute the orientation plane of the scan by computing the main direction of the normal to the slices [2]. But even in the aforementioned method missing slices or extra slices, unidentified attributes and unexpected errors may cause difficulty in detecting the orientation plane of the brain MR scan.There is very little work done to detect the orientation plane of brain MR scans especially using pixel data and deep learning. Hence, with the use of the pixel data and deep learning methods we achieved to train 3 different models to predict the orientation of the brain MR scans.

## 3 Data Preparation

The dataset used in this work comes from The Cancer Imaging Archive (TCGA - LGG) [7]. It was filtered to obtain the subset of images having Image Modality being MR. Two subsets (A and B) of the dataset were downloaded. Subset A consisted of 40 out of the 199 total subjects from the TCGA-LGG dataset which in turn had 551 series. Subset B consisted of 20 out of the remaining 159 subjects, which further gave 299 series. Both subsets A and B were then filtered so that they only consisted of series with sequence T1, T2 or FLAIR. This filtering was done with the help of the series description attribute of the DICOM files, however, due to variation and multiple sequence tagging in the values of series description, 19 out 551 series in subset A and 42 out of 299 series in subset B were manually verified and annotated. Finally, subset A contained 266 series and subset B consisted 125 series. We used subset A for the purposes of training and validation, in the ratio 75:25 respectively. Subset B was used as the test set for model evaluation.

## 4 Extracting ground truth labels

The series were classified into three different labels - Axial, Sagittal and Coronal, which are the standard and most common orientation planes for brain MR scans. Instead of annotating all the data manually, most labels the labels were derived from the Image Orientation Patient (0020,0037) DICOM attribute. The first three values are considered as vector 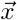 and the other three values are considered as vector 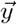, after which the cross product of vectors 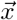 and 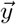 is calculated. If the resultant vector’s *î* component value is 1 then the series is labeled as ‘Axial’, if not then the *ĵ* component is considered. Then, if the *ĵ* component has the value 1 then the series is labeled as ‘Coronal’, if not then we move on to checking the 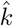 component. Finally, if the 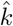 component value is 1 then the series is labeled as ‘Sagittal’ [8]. For each series, the SeriesInstanceUID and its derived orientation plane (ground truth) are stored in a csv file for training and evaluation purposes.

## 5 Methodology

The preprocessing happens at the series level. Each series contains multiple DICOM images (instances). Each DICOM image (instance) is converted to an 8-bit NumPy array of the given size and is processed 4 times, with four different windows each time. Windowing can be understood as a means of manipulating the high amount of voxel values in the DICOM images in order to change the appearance of the image so particular features an image look highlighted [9]. The pixel data of each of these four different windows with different feature extraction are appended in four different lists. Each list containing pixel data of all the DICOM images (instances) with a particular window is called a stack. Hence, we created only four unique stacks with four unique windows, which were then used with different approaches.

### 5.1 The Greyscale MPSM Approach

In MPSM both the M stands for mean, P stands for peak-to-peak which is the range of values (maximum - minimum) along an axis and S stands for standard deviation which is the square root of the average of the squared deviations from the mean. In this approach we use the four stacks. Each stack was processed to find the characteristic pixel data values as NumPy arrays. We create two NumPy arrays using the mean values of pixel data along axis 0 of Stack 1 and 4 respectively, similarly, then we find two other NumPy arrays having peak-to-peak values and standard deviation values of Stack 2 and 3 respectively. These four NumPy arrays were then used to form four new images of size 128×128 pixels each which were then pasted on top of each other on a newly created blank image of size 256×256 pixels. The final image was then stored with it’s filename being SeriesInstanceUID. This is done for each series and the resultant image looks like the following figures -

**Figure 1:**
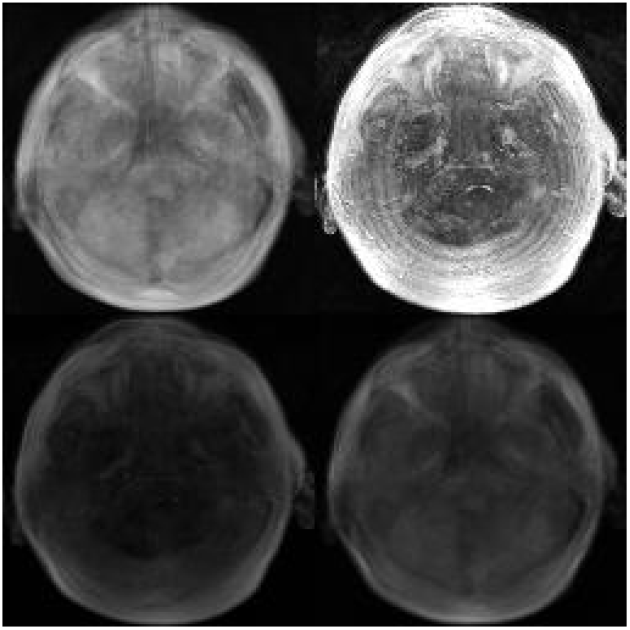
Axial Image

**Figure 2:**
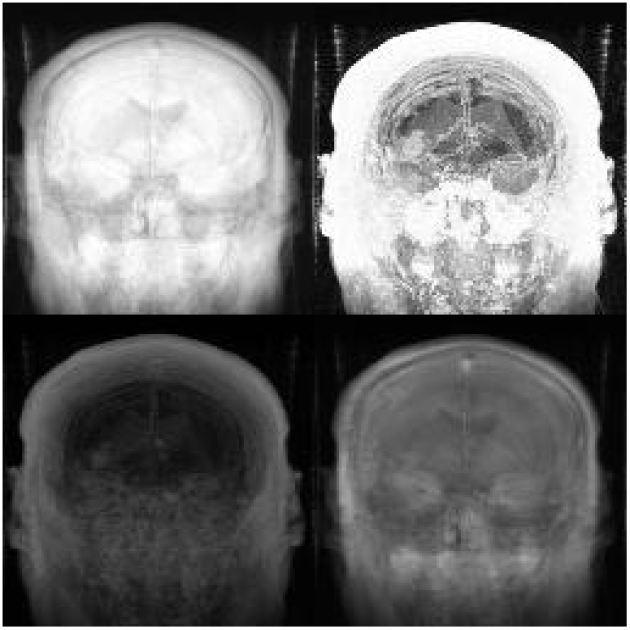
Coronal Image

**Figure 3:**
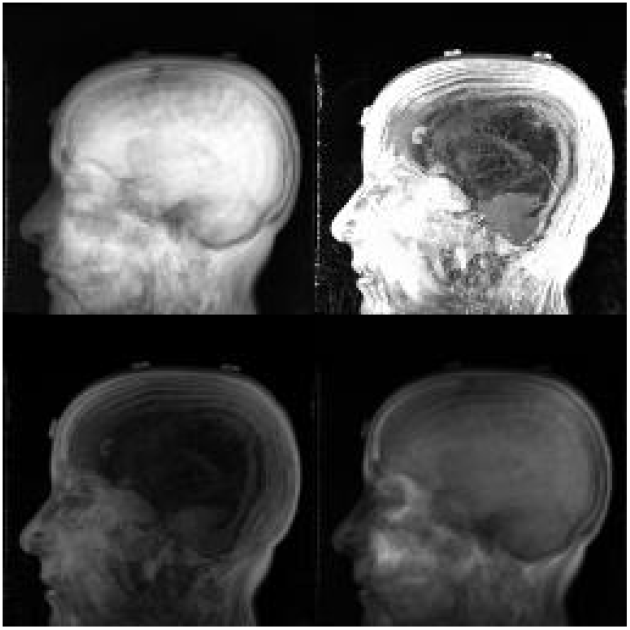
Sagittal Image

### 5.2 The Greyscale 4M Approach

Here the 4M stands for mean, maximum, minimum and mean. Similar to the first approach, we use the above-mentioned four stacks. Each stack was processed to find the characteristic pixel data values as NumPy arrays. We create two NumPy array using the mean values of pixel data along axis 0 of Stack 1 and 4 respectively, similarly, then we create two other NumPy arrays having maximum values and minimum values of Stack 2 and 3 respectively. These four NumPy arrays were then used to form four new images of size 128×128 pixels each which were then pasted on a newly created blank image of size 256×256 pixels. Similar to previous approach the final image was then stored with its filename being SeriesInstanceUID. This is again done for each series and the resultant images look like the following figures -

**Figure 4:**
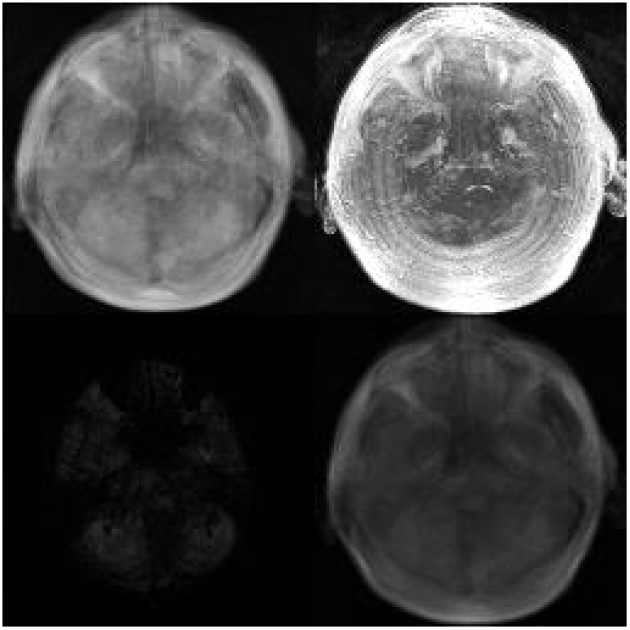
Axial Image

**Figure 5:**
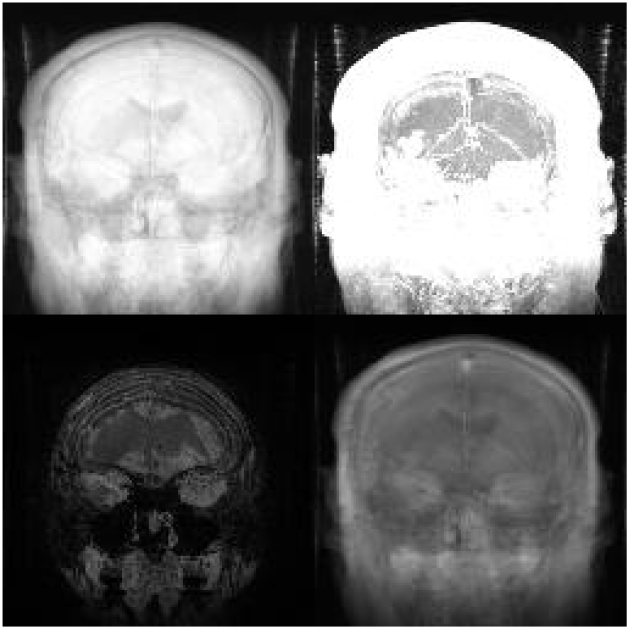
Coronal Image

**Figure 6:**
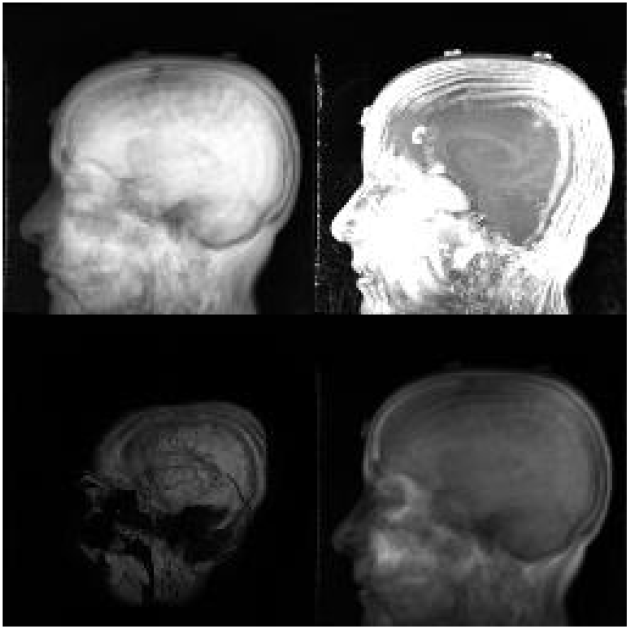
Sagittal Image

### 5.3 The MPSM Approach with both Greyscale and RGB

This approach is similar to Greyscale MPSM approach. In addition to using the greyscale pixel data, we also utilize each channel of the RGB scale. Each stack was again used to find the characteristic pixel data values as NumPy arrays. We created two NumPy arrays having mean values, peak-to-peak values, standard deviation values and again mean values of Stack 1, 2, 3 and 4 respectively in greyscale. Then four new NumPy arrays having the same values as im1, im2, im3 and im4 respectively were created but were converted to the R channel of RGB scale and hence were no longer greyscale. Similarly another four arrays having the same values were converted to the G channel of the RGB scale and NumPy arrays and finally another four NumPy arrays were converted to the B channel of the RGB scale. These sixteen NumPy arrays were used to form sixteen new images of size 64×64 pixels each which were then pasted on a newly created blank image of size 256×256 pixels. Similar to the previous approaches the final image was then stored with its filename being SeriesInstanceUID. Thus, for each series with the help of these 16 resultant NumPy arrays, we create 16 images pasted on a single image, as shown in the following figures -

**Figure 7:**
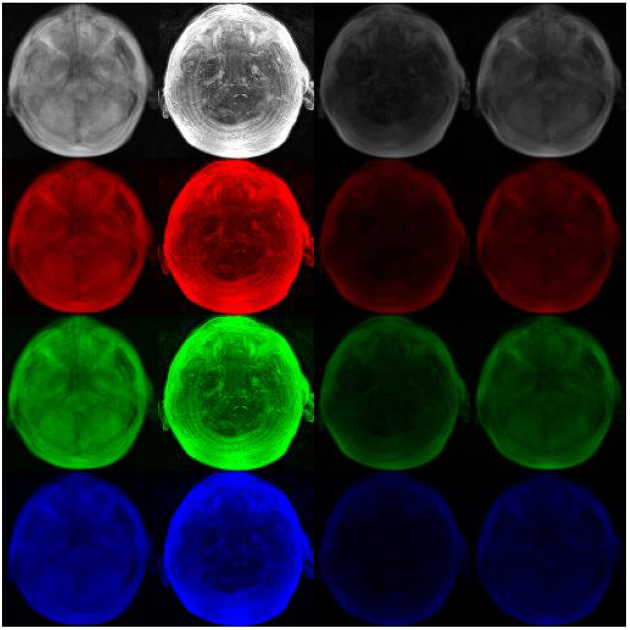
Axial Image

**Figure 8:**
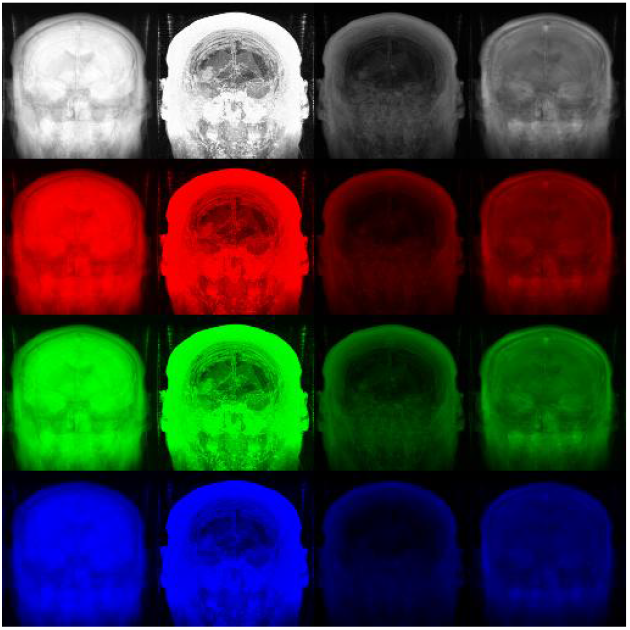
Coronal Image

**Figure 9:**
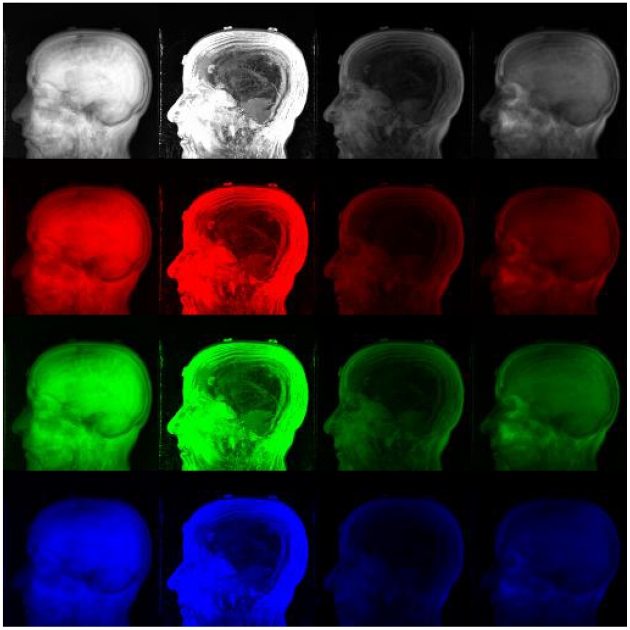
Sagittal Image

## 6 Model

We built a single-label classifier using the FastAI library [10], keeping in mind the ease of use and reproducibility aspects. We also maintain a csv file consisting of all the SeriesInstanceUIDs and their corresponding orientation. We then create a FastAI object that combines the data and a pre-trained convolutional neural network model, in our case squeezenet1_0 [11], and also used transfer learning to fine-tune the pretrained model. We chose squeezenet as it is a small CNN with fewer parameters, hence provides quick inference results with minimal degradation in performance and does not have extravagant computational cost. Squeezenet provides similar accuracy as AlexNet, which is 3 times slower, 500 times bigger and has 50X more parameters than squeezenet [11]. The model was then exported and saved.

## 7 Result

The models were tested on subset B and the following results were obtained -

**Table 1:** Results

| Approach | Testing Accuracy |
| --- | --- |
| Greyscale MPSM | 100 |
| Greyscale 4M | 100 |
| Greyscale + RGB MPSM | 100 |

## 8 Future Work

By using pixel data, we were able to develop a deep learning model that precisely identifies the orientation (Axial/Sagittal/Coronal) of brain MR scans. The next step is to develop a deep learning model that will predict the orientation of brain MR scans (T1/T2/FLAIR) which we can then integrate with the orientation detection model. We will be open-sourcing our method and will be updating this manuscript with further details.

## Data Availability

We used publicly available data from the TCGA website.

https://wiki.cancerimagingarchive.net/display/Public/TCGA-LGG

## Notes

### Competing Interest Statement

The authors have declared no competing interest.

### Funding Statement

None

